# Identifying falls risk using wearables data in older adults: an observational cohort study

**DOI:** 10.1101/2025.11.27.25341162

**Authors:** Atul Anand, Marie Guglielminetti, Greig Fotheringham, Laurie Auld, Jo Gordon, Adrian Smales, Dawn A Skelton, Alexandra Melling, Garrett Sprague

## Abstract

**Background:** Falls are a major cause of morbidity in older adults. Low-cost wearable devices track potential falls risk factors, but adoption in older adults remains uncertain.

**Methods:** We conducted a 6-month prospective observational study in community-dwelling adults who self-reported a recent fall or were deemed at increased risk. Participants were given a wrist-worn wearable device (Fitbit, Garmin or Polar), synced with a smartphone application (Smplicare app) to collect additional information by questionnaires, including self-reported falls. We analysed adherence wearing the devices, and studied step count and sleep data in relation to falls.

**Results:** Of 284 people (74.2±9.0 years, 68% women) in the study, 266 (94%) provided at least 7 days of data, with 196 (76%) engaged on at least half of study days. Engagement did not differ by self-reported technology confidence. There were 81 (30%) people who reported a fall during follow-up, but only 5 (6%) resulted in hospital attendance. Each additional hour of average sleep was associated with a 24% reduction in falls risk (HR 0.76, 95% CI 0.63 to 0.92), but in multivariable models only carer support (aHR 3.47, 95% CI 1.46 to 8.26) and incontinence (aHR 2.26, 95% CI 1.34 to 3.82) remained independently associated with falls. No changes in step or sleep patterns were noted after falls, but there was high individual heterogeneity.

**Conclusion:** Wearable adoption, risk factor identification and digital self-reporting of falls is feasible in older adults using low-cost commercial technology. The importance of simple wearable measures like sleep for fall risk were outweighed by markers of frailty. Future research should understand how these granular wearable data could add to proactive falls risk assessment.

## INTRODUCTION

Injurious falls are a major frailty marker in older adults, with around 1 in 5 older people dying within a year of suffering a fragility fracture.[1, 2] Over 80,000 older people sustain a hip fracture each year in the UK, costing the NHS and social care over £2 billion,[3] rising to over £4 billion if other fragility fractures are included. Beyond mortality, falls negatively affect wellbeing in older adults, frequently triggering a precipitous cycle of decline in mobility related to fear of further falls, subsequent muscle loss from inactivity and increasing dependence.[4] Healthcare systems are striving to keep people living healthy independent lives for as long as possible – to extend ‘healthspan’ rather than just lifespan – and falls prevention represents a critical target.

Longitudinal tracking of health data, including by wearable devices, has potential to support a shift to proactive intervention in those at risk of an injurious fall. The recent World Falls Guideline for prevention and management found insufficient evidence to support the use of wearables,[5] but did note some evidence of improved participation within structured exercise programmes when simple wearable devices were used.[6] Frequently, wearables data has focused on step counts, but commercial wrist-worn ‘smart watches’ are more sophisticated in their tracking abilities, including metrics like patterns of sleep and heart rate variability.[7] Adoption across the population has dramatically grown as cheaper products have become increasingly ubiquitous.[8] However, use and acceptability of these devices amongst older adults remains an area of uncertainty. Prior small studies have reported mixed results in this area, with notable challenges in those with declining cognition and one small study reporting devices worn on fewer than a third of days for a third of participants.[9-11]

The incidence of falls is misunderstood if only relying on healthcare contacts or studies of fractures following falls. It has long been recognised that most community falls do not result in a healthcare interaction,[12] and these potential opportunities for intervention are usually missed outside of longitudinal research studies. The most recent NICE guidelines for falls assessment and prevention recommends people aged 50 and over at higher risk are supported with evidence-based interventions, and one of their key recommendations for research is identifying how accurate wearable technologies are at identifying risk in real world settings.[13] Wearable devices are increasingly recognised as a means of detecting falls and simple digital technologies such as smartphone applications provide more opportunities for self-reporting of falls or emerging risk factors.[14]

Our aim was to assess the adherence and outputs of commercial wearable devices and digital health data collection in relation to risk of fall for older people.

## METHODS

### Study population and setting

We conducted a prospective observational study of self-reported falls over a 6-month period, in community dwelling adults aged >55 years old in the United Kingdom. Participants were eligible if they self-reported a recent fall (within the prior 12 months) or were deemed to be at an increased falls risk either by self-report or referral from a study partner, including community falls prevention programmes and housing associations. Participants were not eligible if unable to provide informed consent, or if unwilling to use a study smartphone/tablet device (to access the data collection application developed by Smplicare) or a wrist wearable device.

### Wearables and enrollment

Study participants were provided with a wrist wearable device without cost and a tablet device if required. Included wearable devices were midrange commercially available products from Fitbit, Garmin and Polar with the ability to capture activity and sleep measures. The informed consent process was undertaken in-person, with a researcher then helping the study participant to register their wearable device with their mobile application (essential to allow user data to flow into a study database) and troubleshoot any device or connection issues. Enrollment was considered complete after device syncing of any wearable and questionnaire data with the study database.

### Data collection

Baseline height and weight were obtained at enrollment to calculate a body mass index (BMI) and hand grip strength was recorded as the best of three trials using a Jamar dynamometer on the dominant arm from a seated position. Baseline data for demographics, function, disabilities and morbidities were collected by questionnaires delivered through the dedicated study smartphone application. An additional baseline question established an individual’s technology confidence level on a scale from uncomfortable to very comfortable. Participants had the ability to report a fall via the application at any time, with a further prompt by weekly questionnaires. Secondary questions after reporting a fall included determining if this resulted in hospitalisation. Wearable device data for step counts and sleep hours were transferred to the study database at each sync of the participant’s data with the mobile application.

### Outcomes

The primary outcome for this analysis was the first fall within the study period, as self-reported by the study participant. Secondary outcomes included adherence to the wearable device and change in step count or sleep hours before and after a fall. Device adherence was determined by the number of participation days, defined as a day in which either step or sleep count data (non-zero values) were successfully transferred into the study database.

### Statistical analysis

The analysis population excluded indiviudals who provided less than 7 days of wearables data across the entire study period. Baseline differences between people who went onto to register a fall (‘fallers’) and those who did not (‘non-fallers’) were compared, reporting frequencies (percentage) for categorical variables, and mean (standard deviation) or median (interquartile range) for continuous variables depending on distribution. Cox proportional hazard models were constructed for an outcome of first self-reported fall, including days to that fall within the survival model. Following univariable analysis of relevant covariates, three multivariable models were created, first combining demographics with morbidity count, then demographics with wearables data, and finally including demographics, morbidity count and wearables data in a single fully adjusted model. Outputs were reported as the hazard ratio for a self-reported fall with 95% confidence intervals (95% CI). Kaplan-Meier plots for survival free from fall were created stratified by tertiles of average step count and sleep duration during the first 30 days of the study. For these analyses, fallers within this 30-day observation period were excluded. To assess change in individual participant level activity around the time of a fall, a 14-day window was created before and after the first reported fall in each participant, and the average step count and sleep hours were calculated for these periods. Participants were excluded if reporting less than 7 days data either before or after their fall. For comparison to non-fallers, a 14-day window was assessed in the same manner before and after their median study participation day.

### Ethics

The study and its protocol were reviewed by the Edinburgh Medical School Research Ethics Committee (EMREC reference 22-EMREC-045). Informed consent was obtained from all participants. All study procedures were completed by Smplicare staff trained in research practice, following the approved protocol.

## RESULTS

Of the 284 people (mean age 74.2 ± 9.0 years, 68% women) enrolled in the study, 266 (94%) provided at least 7 days of wearables data across the 6-month study period and so formed the analysis population. Of this group, 81 (30%) reported at least one fall during 6 months of follow-up, but only 5 (6%) of these events resulted in hospital attendance or admission. **Table 1** shows baseline characteristics of the analysis population stratified by fall status. People who reported a fall during the study were more likely to report any carer support (23% *vs* 8% without a fall, p=0.002) and continence issues at baseline (58% *vs* 33%, p<0.001), but a history of a fall in the prior 6 months was not different between groups (84% vs 74%, p=0.09). Fallers and non-fallers during the study were similar in age, sex, ethnicity, self-rated health and morbidities recorded at baseline. On average, people who fell during the study trended towards more physical activity recorded on their wearable device (median 6006 steps per day vs 4808 in non-fallers, p=0.07). Fallers recorded 24 fewer sleep minutes per night compared to non-fallers (mean 6.8 hours *vs* 7.2 hours respectively, p=0.01). These differences between groups remained stable across the study period (**Supplementary Figure S1**).

**Table 1.** Baseline characteristics stratified by self-reported fall status at any point during study period. All values are counts (%) unless otherwise stated. Activity data represented from across study period.

|  | All | Faller | Non-Faller | p-value |
| --- | --- | --- | --- | --- |
| N | 266 | 81 | 185 |  |
| ***Demographics*** |  |  |  |  |
| Age, mean (SD) | 74.4 (8.9) | 74.1 (8.8) | 74.5 (9.0) | 0.75 |
| Men | 83 (31) | 26 (32) | 57 (31) | 0.95 |
| Ethnicity |  |  |  | 0.73 |
| White | 234 (88) | 72 (89) | 162 (88) |  |
| Asian | 13 (5) | 3 (4) | 10 (5) |  |
| Black | 12 (5) | 3 (4) | 9 (5) |  |
| Other or Unknown | 7 (3) | 3 (4) | 4 (2) |  |
| ***Measures*** |  |  |  |  |
| BMI, kg/m<sup>2</sup>, mean (SD) | 27.0 (8.2) | 26.8 (8.4) | 27.2 (8.1) | 0.75 |
| Grip Strength, kg, mean (SD) | 24.7 (10.6) | 24.4 (11.4) | 24.8 (10.2) | 0.77 |
| ***Questionnaires*** |  |  |  |  |
| Carer support | 32 (12) | 18 (23) | 14 (8) | **0.002** |
| Fall in last 6 months | 204 (77) | 68 (84) | 136 (74) | 0.09 |
| Any incontinence | 100 (41) | 45 (58) | 55 (33) | **<0.001** |
| Use of walking aid | 93 (38) | 32 (41) | 61 (37) | 0.67 |
| Instrumental ADL dependency | 158 (65) | 54 (69) | 104 (63) | 0.42 |
| Basic ADL dependency | 51 (21) | 20 (26) | 31 (19) | 0.28 |
| Self-rated health |  |  |  | 0.43 |
| Excellent | 8 (3) | 3 (4) | 5 (3) |  |
| Very good | 56 (23) | 14 (18) | 42 (26) |  |
| Good | 104 (43) | 31 (40) | 73 (44) |  |
| Fair | 65 (27) | 26 (33) | 39 (24) |  |
| Poor | 10 (4) | 4 (5) | 6 (4) |  |
| ***Morbidities\**** |  |  |  |  |
| Total count, median [Q1, Q3] | 3 [2, 4] | 3 [2, 4] | 3 [2, 4] | 0.42 |
| Atrial fibrillation | 25 (10) | 8 (10) | 17 (10) | 1 |
| Hypertension | 135 (56) | 40 (51) | 95 (58) | 0.40 |
| Myocardial Infarction | 21 (9) | 10 (13) | 11 (7) | 0.18 |
| Stroke | 36 (15) | 16 (21) | 20 (12) | 0.13 |
| Diabetes | 46 (19) | 16 (21) | 30 (18) | 0.81 |
| Asthma | 46 (19) | 14 (18) | 32 (20) | 0.91 |
| COPD | 26 (11) | 6 (8) | 20 (12) | 0.40 |
| Chronic Kidney Disease | 20 (8) | 6 (8) | 14 (9) | 1 |
| Depression | 62 (26) | 23 (30) | 39 (24) | 0.43 |
| Dementia | 8 (3) | 5 (6) | 3 (2) | 0.14 |
| Arthritis | 147 (61) | 53 (68) | 94 (57) | 0.15 |
| Osteoporosis | 70 (29) | 23 (30) | 47 (29) | 1 |
| Hip fracture | 16 (7) | 4 (5) | 12 (7) | 0.72 |
| ***Activity data*** |  |  |  |  |
| Daily steps, median [Q1, Q3] | 4882 [2574, 7458] | 6006 [2789, 8368] | 4808 [2450, 7042] | 0.07 |
| Daily sleep, hours | 7.0 (1.2) | 6.8 (1.4) | 7.2 (1.1) | **0.01** |

ADL = Activities of Daily Living. Basic refers to assistance required with washing, dressing, feeding or toileting. Instrumental refers to assistance required with shopping, cleaning house, cooking, managing finances and laundry. \*Reported in 244 (92%) of participants returning morbidities data

### Wearable device adherence

There was a gradual decline in the number of study participants contributing wearables data on each study day, as shown in **Figure 1A**. Data was available on at least half of the study days in 196/266 (76%) people and for three quarters of study days in 163/266 (61%). The proportion of active users providing valid step count data was stable throughout the study (**Figure 1B**) with overall availability for 95.2 ± 2.0% of users on days in which their device was active. A similar stable pattern was observed for returns of sleep data (**Figure 1C**) but for a lower proportion of users (81.8 ± 5.1%). Median days of data contribution did not differ by baseline self-reported technology confidence level at 155, 168 and 167 days for ‘less than comfortable’, ‘comfortable’ and ‘very comfortable’ respectively (p=0.47, **Supplementary Figure S2**).

**Figure 1.**
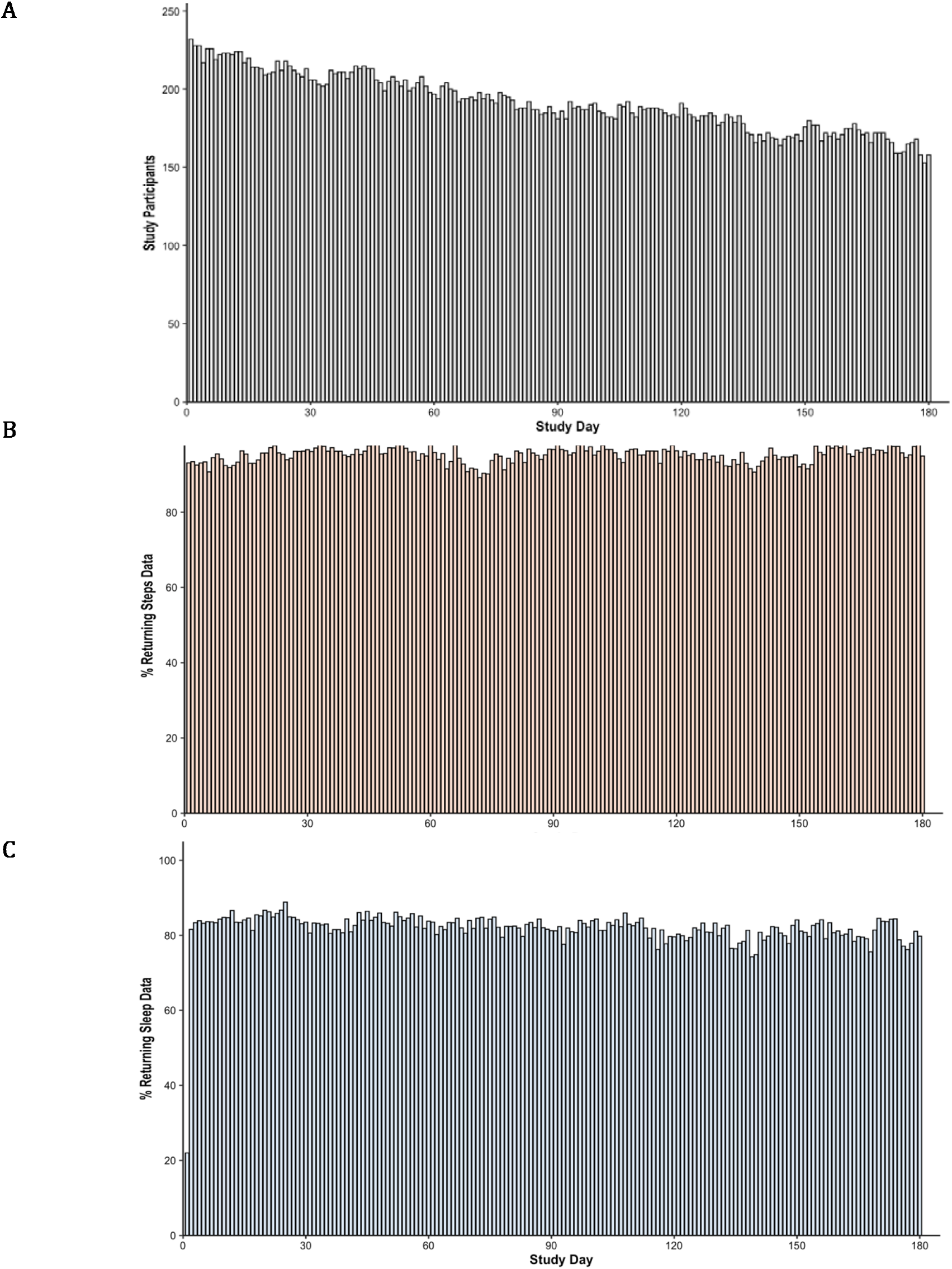
Wearable device adherence. Charts show a count of the number of study participants contributing wearables data on each study day (A), and proportion of active participants on those days with valid step count (B) and sleep data (C).

### Predictors of falls

Carer support and a history of incontinence were both strongly associated with a greater than doubling of the risk of falling during the study period in univariable Cox proportional hazard models (**Table 2**). The relationship for carer support strengthened in multivariable models including demographics, disability, falls history, morbidities and wearables data, reaching a three-fold increased risk in the fully adjusted model (HR 3.47, 95% CI 1.46 to 8.26). In univariate associations, each increased hour of average sleep time was associated with a 24% reduction in falls risk (HR 0.76, 95% CI 0.63 to 0.92), but this relationship was not retained after adjustment for covariates.

**Table 2.** Univariable and multivariable Cox proportional hazard models for an outcome of first fall during the study period. Model 1 includes baseline demographics, questionnaire responses and morbidity count data (n=229). Model 2 includes demographics, questionnaire responses and wearables data (n=209) and Model 3 includes all variables (n=207). All figures are hazard ratios with 95% confidence intervals and are emboldened when significant at p<0.05.

|  | Univariable | Model 1 | Model 2 | Model 3 |
| --- | --- | --- | --- | --- |
| N | – | 229 | 209 | 207 |
| Age, per year | 1.00 (0.97–1.02) | 0.99 (0.96–1.02) | 1.00 (0.97–1.03) | 1.00 (0.97–1.03) |
| Male sex | 1.10 (0.69–1.75) | 1.34 (0.81–2.21) | 1.18 (0.68–2.03) | 1.23 (0.71–2.13) |
| BMI, per kg/m<sup>2</sup> | 1.00 (0.97–1.03) | 1.00 (0.97–1.03) | 1.01 (0.98–1.05) | 1.01 (0.97–1.04) |
| Carer support | **2.78 (1.64–4.70)** | **3.79 (1.80–7.98)** | **3.36 (1.43–7.90)** | **3.47 (1.46–8.26)** |
| Any incontinence | **2.29 (1.46–3.58)** | **2.42 (1.50–3.92)** | **2.27 (1.35–3.82)** | **2.26 (1.34–3.82)** |
| Use of walking aid | 1.16 (0.74–1.83) | 0.75 (0.44–1.30) | 0.84 (0.44–1.60) | 0.77 (0.40–1.47) |
| Instrumental ADL dependency | 1.30 (0.81–2.11) | 1.05 (0.61–1.83) | 1.09 (0.61–1.96) | 1.04 (0.57–1.87) |
| Basic ADL dependency | 1.48 (0.89–2.47) | 0.71 (0.34–1.48) | 0.86 (0.39–1.90) | 0.79 (0.35–1.79) |
| Fall in last 6 months | 1.69 (0.93–3.06) | 1.40 (0.74–2.64) | 1.38 (0.71–2.69) | 1.32 (0.68–2.58) |
| Morbidities, per condition | 1.09 (0.97–1.22) | 1.07 (0.93–1.23) |  | 1.10 (0.95–1.27) |
| Daily steps, per 1000 | 1.04 (0.98–1.11) |  | 1.06 (0.97–1.15) | 1.06 (0.97–1.15) |
| Daily sleep, per hour | **0.76 (0.63–0.92)** |  | 0.88 (0.72–1.07) | 0.90 (0.73–1.09) |

### Wearable activity in relation to falls

Figure 2. shows survival free of falls by tertile of average step count and sleep time recorded in the first 30 days of the study, excluding the 27 study participants who recorded a fall in this period. There were 54 participants with a first fall during the remaining 150 days of observation. No relationship was observed between average step count and falls (p=0.5, **Figure 2A**). However, falls risk was greatest in the tertile with shortest average sleep time (<6.8 hours) and lowest in the middle tertile for sleep (6.8 to 7.5 hours) with a difference between groups (p=0.02, **Figure 2B**).

**Figure 2.**
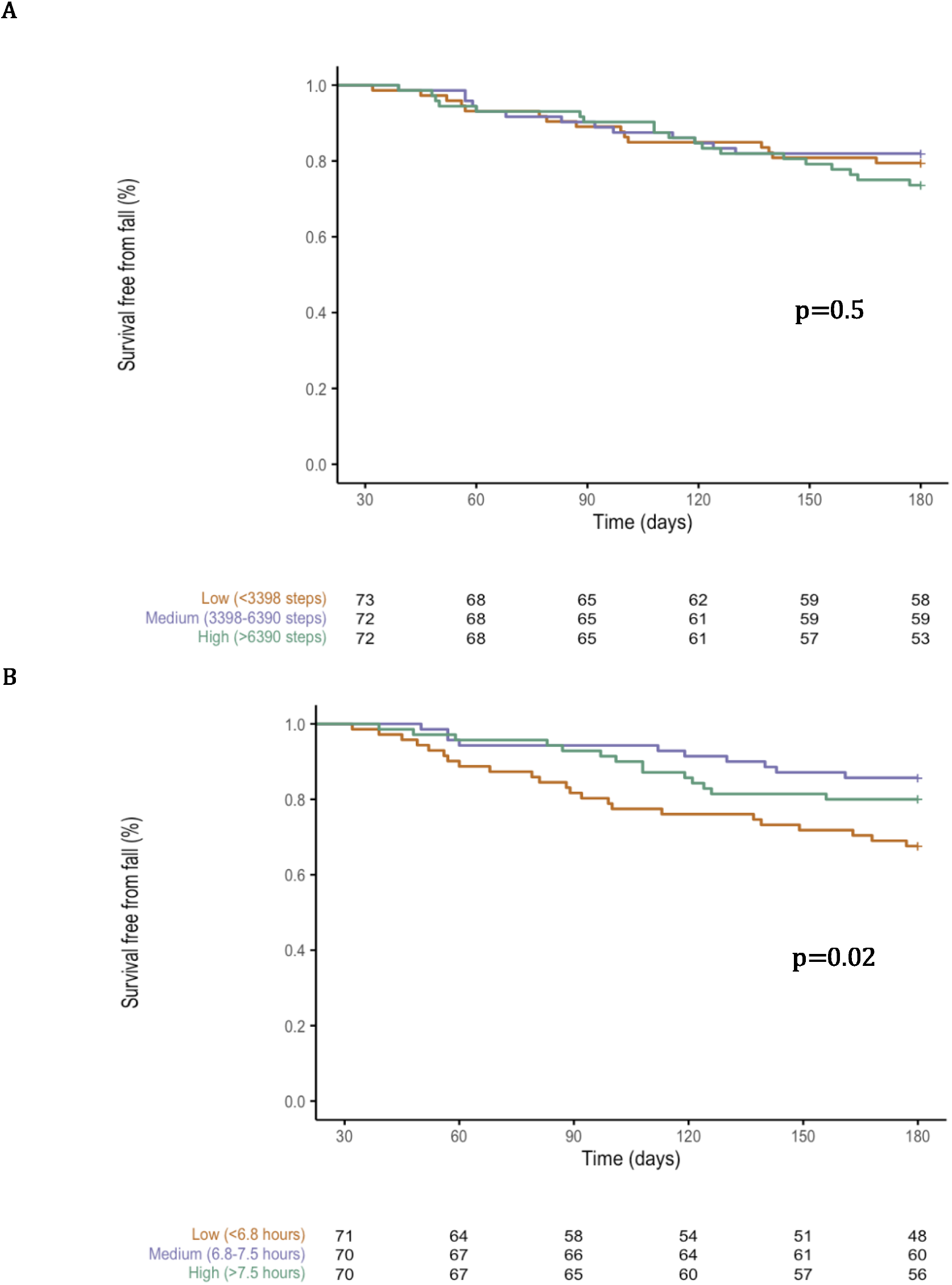
Kaplan-Meier plots for survival free from falls between study day 30-180, stratified by tertile of average daily step count (A) and sleep hours (B) over the first 30 days of observation.

Considering individual wearables data, there was no change in step count in the 14 days before and after a first fall (change +110 steps per day after fall, 95% CI −294 to +514 steps). Similarly in non-fallers using the 14 days either side of their median study day, no change was observed (−110 steps per day, 95% CI −475 to +255). Comparing between groups there was no difference in this change (p=0.42). For sleep duration there was no change observed after either a first reported fall (−0.04 hours, 95% CI −0.30 to +0.21) or the median study day in non-fallers (−0.06 hours, 95% CI −0.18 to +0.06). Between group differences were again non-significant (p=0.92). However, wide heterogeneity in step and sleep patterns were observed in both fallers and non-fallers (**Figure 3**).

**Figure 3.**
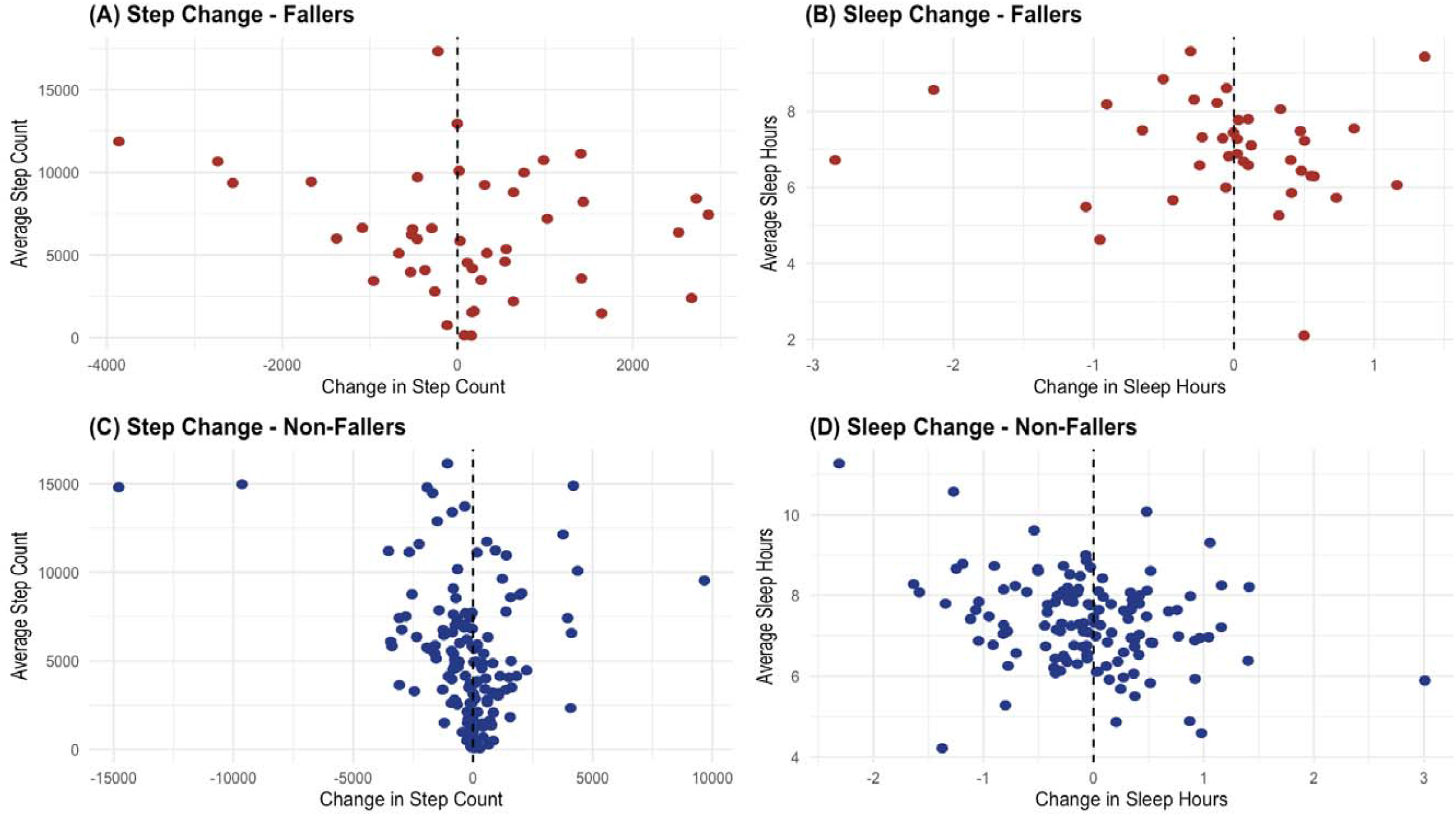
Change in daily step count (A and C) and sleep duration (B and D) in the 14 days after a fall or the median study participation day in non-fallers. Points are plotted against the average daily step count or sleep hours in the 14 days prior to the fall or median study day. Fallers (A and B) are shown in red, while non-fallers (C and D) are shown in blue.

## DISCUSSION

This prospective observational cohort study in people with a history or high risk of falls showed good uptake and adherence with wearables devices and digital falls recording over a 6-month period. Participation levels were similar regardless of confidence with technology at the start of the study. A third of those included reported a fall within the study period, confirming the high risk of those enrolled, but only 1 in 20 self-reported falls resulted in a recorded hospitalisation. Wearables data identified an association between lower sleep and falls risk, but this was outweighed in multivariable models by other measures of overall frailty, notably the requirement for carer support and incontinence. Taken together, our findings show potential for wearable technology and digital health recording to inform proactive care for older people at high risk of falls. High heterogeneity in the highly granular step count and sleep information suggests that more advanced data methods might improve risk prediction for falls, but there remains uncertainty in the role and response of health and social care providers to alerting change in these areas.

Our study suggests that most participants successfully wore their device and synced data for most days of the study. These findings are in line with a recent study of 175 older adults enrolled in the UCSF Longitudinal Brain Aging Study using a similar commercial FitBit device, although that protocol only required daytime wearing over 30 days.[9] Studies in this area are often similarly short in observation period, providing limited insights into sustained use and effects of wearables. A systematic review found only 8 of 117 included studies reporting outcomes beyond 6 months, with the small increase in physical activity observed amongst wearable users slightly diminishing over time.[15] The evidence in people living with dementia or mild cognitive impairment is more limited and merits wider consideration of appropriate technology design that might not be a priority for commercial manufacturers.[11] Collaborative co-design involving older people and practitioners in setting goals has been shown to improve adherence with wearables.[16]

The wider area of digital technology in older adults is growing at pace; a recent systematic search found hundreds of smartphone applications targeted at improving physical activity in this group, but only one with any form of published evidence.[17] As the cost of commercial technology – both smartphones and wearables – continues to fall, the barrier to market entry is ever diminishing. This offers opportunities for cost-effective and rapid scale-up of care pathways informed by these tools, but provides an onus on health and care systems to evaluate the efficacy of these new approaches. Given the growth in adoption of wearables, there is an inevitability that device use will become more common in our older population and clinicians will have new opportunities to review physical activity and other metrics outside of direct measurement in health settings.[18] Integration of these highly granular data is complex, with even ‘basic’ commercial wearables now collecting multiple datapoints multiple times per minute. We adopted a simplistic aggregation of daily data in our study, but machine learning and artificial intelligence approaches might gain more informative insights on individual risk from these data. This area is still in its infancy for falls prediction but has potential to change our approach to proactive falls reviews in the future.[19]

Our observation of an association between fewer hours of sleep and falls has been noted in some prior literature. A systematic review and meta-analysis of 7 observational studies suggested a ‘U-shaped’ association similar to our findings, with increased falls risk in those with relatively short and long sleep times.[20] However, all studies used self-reported sleep and variability in reporting of these measures has been noted in other work.[21] Low sleep time has been associated with numerous harmful health outcomes including cardiovascular diseases and cancers.[22] Tracking sleep from wearable devices might provide objective measurement to understand risks better, particularly when using longitudinal data from the same individual over time.

It is striking that only 6% of first self-reported falls in our study resulted in hospitalisation. A longitudinal study of older people published 50 years ago, noted that most falls in the community did not result in detectable injury, and that “morbidity was negligible” in over 80% of falls.[12] However, this and many other studies have reported the clustering of falls amongst individuals who go on to experience injury. Decades later, it may be argued that we have not successfully spread health or care interventions that substantially mitigate these risks, nor developed the systematic responses to the ‘minor’ fall that might herald a coming injurious episode. The recent World Falls Guideline and updated NICE falls recommendations go some way to acknowledge this challenge and offer a unifying set of evidence-based guidelines.[5] [13] While there is currently insufficient evidence to support inclusion of wearables data in risk assessment, these guidelines promote proactive and opportunistic case-finding and physical activity, both of which might benefit from wearables and digital recording of health and falls risk factors. Others have already attempted to add nuance to recommended falls risk assessments using additional information from physical activity measures, to stratify risk and support triage into more manageably sized groups.[23] However, as with all technological advances, a major barrier remains the integration of new data into complex digital health and social care systems, including appropriate information sharing between professionals across sectors.[24]

Our study has several strengths. We successfully enrolled people at demonstrably high risk of falls and showed our approach to wearables and data collection was feasible within both home settings and community falls programmes. The study population had high levels of multimorbidity and the majority had some dependencies in activities of daily living. However, we acknowledge limitations in this work. Average physical activity levels were high in the study population, suggesting limited generalisability to the frailest populations. We used three different commercial wearable devices and therefore interpretability of readings between manufacturers might be limited. For this reason, we only focused on core measures of step count and sleep hours for this analysis, where the risk of significant differences between devices were thought to be less likely. Finally, although self-report of falls was high, we acknowledge that even this approach might underestimate the true burden of falls.[25] Spread and adoption of these tools is only likely to increase in our ageing population. Future research should focus on understanding the full potential of these highly granular data for informing falls risk assessment within integrated care pathways.

## Conclusion

Receipt of care and incontinence were predictive of a future fall and we have shown that these features can be self-reported by this cohort of older adults using a simple digital application. Lower sleep duration, collected from wearables, was able to highlight a near 3 fold increased risk of fall, while simple step counts did not add to understanding of risk for this cohort. Our study shows the feasibility for low-cost commercially available wearables and digital health technologies to support the identification of falls risk factors and recording of falls events.

## Data Availability

All data produced in the present study are available upon reasonable request to the authors.

## ACKNOWLEDGEMENTS

We would like to thank all the participants in this study for their dedication, and more than 20 cross-sector partners who supported our recruitment efforts.

