## Supplement for "Identifying falls risk using wearables data in older adults: an observational cohort study"

Atul Anand^1^, Marie Guglielminetti^2^, Greig Fotheringham^2^, Laurie Auld^1^, Jo Gordon^3^,

Adrian Smales^2^,Dawn A Skelton^4^, Alexandra Melling^2^, Garrett Sprague^2^

**Figure S1.** Averaged step count (A) and sleep (B) in 30-day blocks of study time, stratified by participants who fell or did not fall at any point during the study period.

**A**

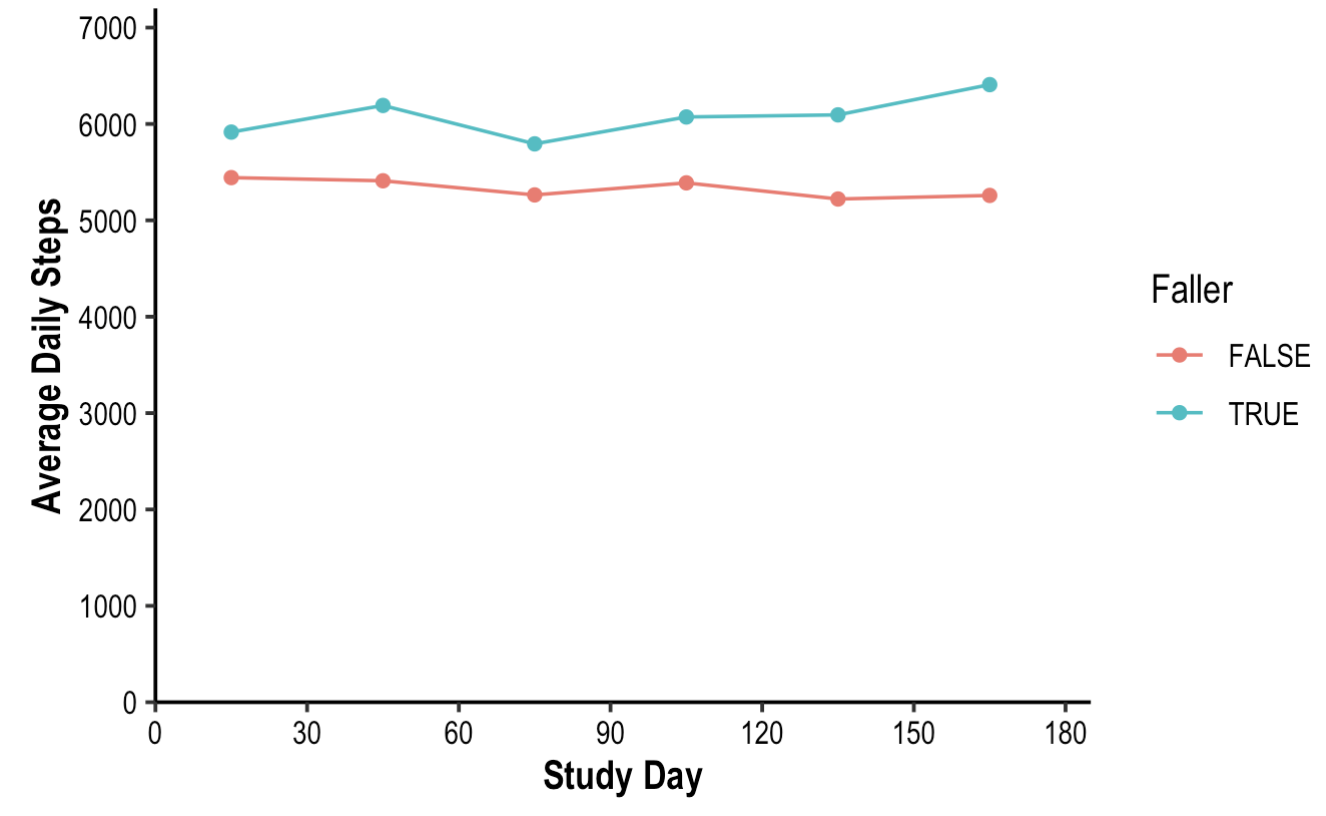

**B**

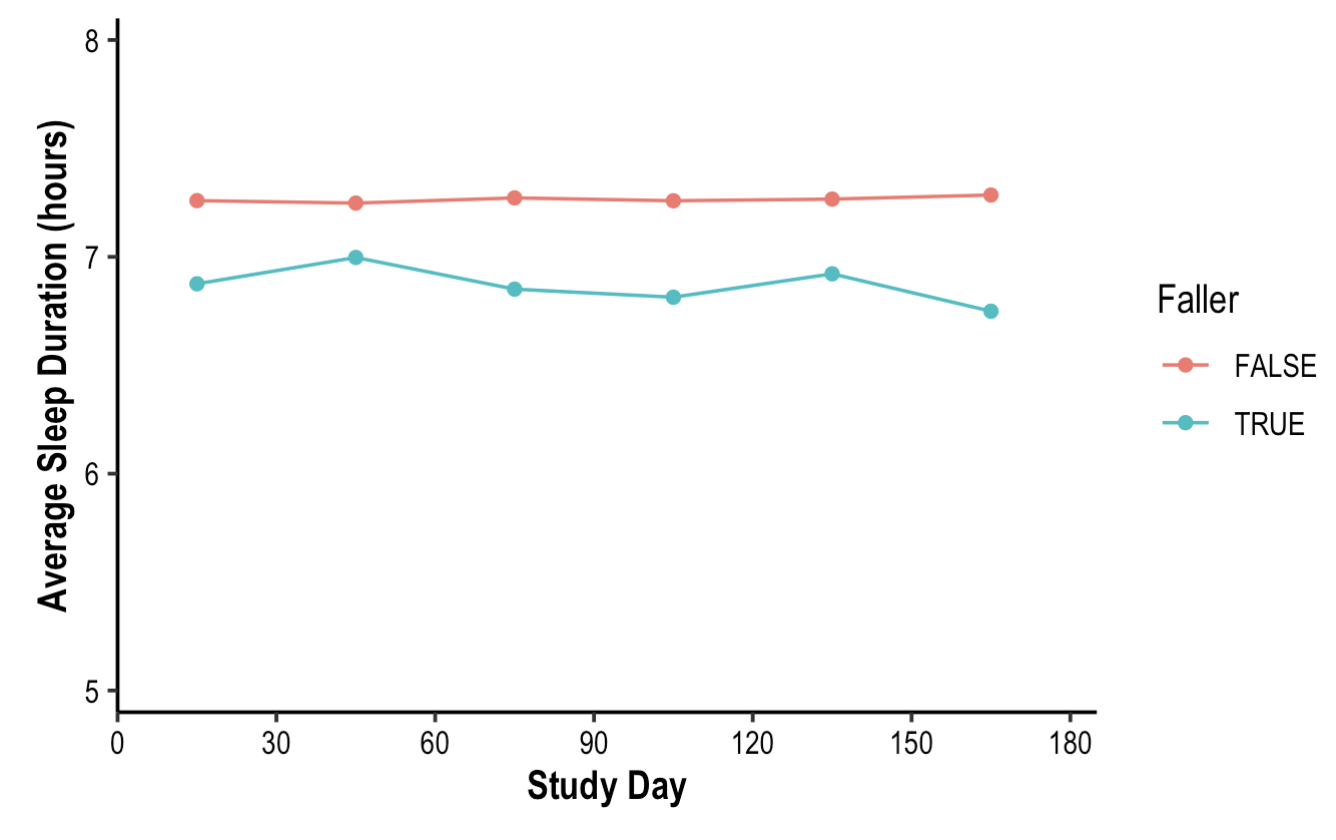

**Figure S2.** Days of participation by technology comfort level.

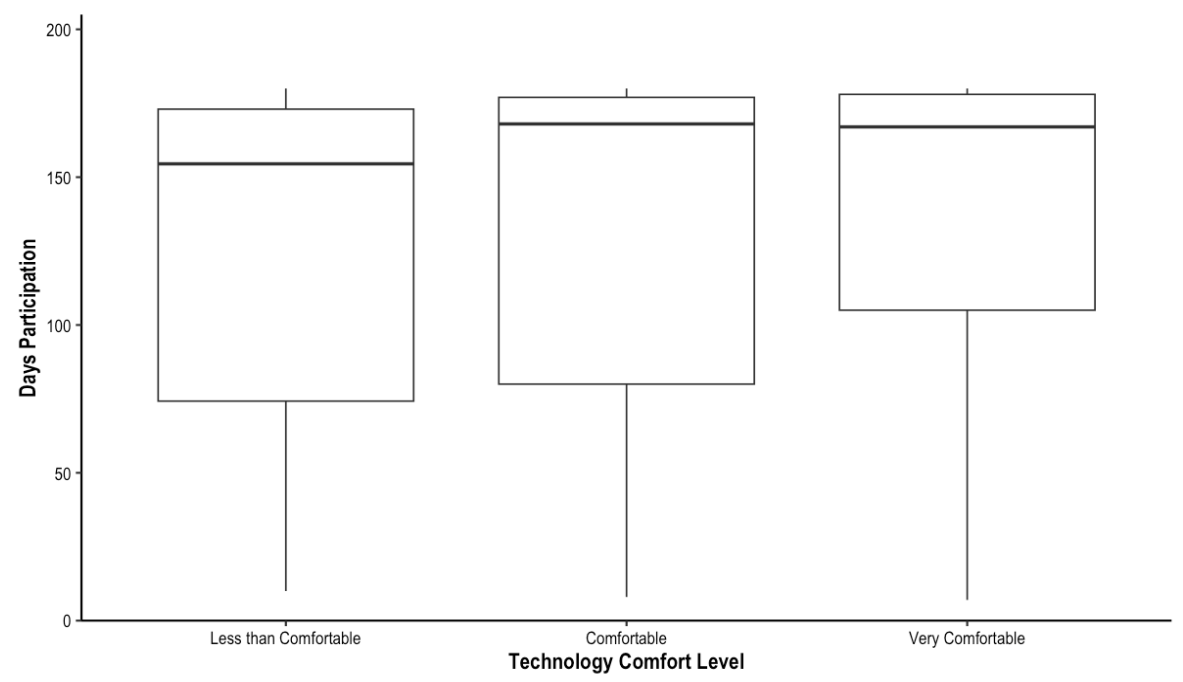
